# Maximising pain services for frail older adults, the views of healthcare professionals and commissioners: findings from the Pain in Older People with Frailty (POPPY) study

**DOI:** 10.1101/2025.09.24.25336537

**Authors:** Alan Wright, Deborah Antcliff, Nicky Kime, Nicola Harrison, Rahena Mossabir, Asim Suleman, Anne Forster, Lesley Brown

## Abstract

**Background:** Chronic pain is common among older adults with frailty and its management often remains suboptimal, despite evidence for the benefits of biopsychosocial treatment approaches being found for other populations. The Pain in Older People with Frailty Study (POPPY) was a four-phase study that aimed to develop a service model for pain management for this population to enable them to better manage their pain and reduce its impact on their lives. The aim of Phase 3 of the POPPY study was to understand the views of those delivering and commissioning services relating to older adults’ engagement in services and how pain services could be maximised to meet their needs.

**Methods:** We used in-depth semi-structured interviews with healthcare professionals (HCPs) and commissioners to explore: 1) perceptions of opportunities and barriers to including and managing older adults in pain services within different contexts, and 2) how to maximise support for this population in community, primary care, secondary care, and tertiary pain services. A thematic approach was used to analyse the data.

**Findings:** We recruited participants from 9 pain and 2 generic community services in the north, southeast and west of England. Services were in community, primary care, secondary care, and tertiary settings. We interviewed 42 HCPs including clinicians, psychologists, allied health professionals, nurses, social prescribers, service managers, and health/wellbeing coaches. We also interviewed 2 service commissioners. Most participants recognised that older adults living with frailty and pain often shared characteristics relating to their physical health, life experience and social circumstances which shaped their engagement in pain services. Generally, participants perceived there to be reduced engagement in pain services among older adults with frailty. Factors that were likely to improve the management of pain in the older population both within pain and non-pain services were also identified.

**Conclusions:** For pain services to meet the needs of older adults with frailty, it is essential for them to be responsive to the specific needs of this population, adapting both the content and delivery of interventions accordingly.

**Trial registration:** research registry7169/ IRAS project ID: 310174

## Background

Chronic or persistent pain, lasting for at least 3 months’ duration becomes increasingly prevalent with age, and is common in older adults due to the increasing likelihood of arthritis and other diseases in this age group (1). Prevalence of chronic pain globally among older adults ranges from 24% to 76% (1, 2). Pain in older adults is associated with disability from reduced mobility, restricted activity, falls, depression, anxiety, impaired sleep, and social isolation (3). The scale of the problem is set to increase in line with the ageing population; nearly a quarter of the UK population are now over 65 years of age, and the incidence of chronic pain is greater in the older demographic (4). Unfortunately, pain in this age group is often under-recognised and under-treated (5). Whilst the evidence base informing how best to manage pain in older adults is limited (3), it is suggested that addressing pain in older adults often requires a different approach compared to younger individuals due to concomitant multimorbidity, polypharmacy, and frailty (5).

Frailty is characterised by age-related decline across multiple physiological systems and vulnerability to disproportionate changes in health or outcomes, for example, falls, disability, or admission to hospital after relatively trivial health events, such as minor infection (6). Frailty affects approximately 12% of people aged 65 years and over, and approximately one third of people aged 80 years and above (7) and is considered a condition requiring long-term strategies and interventions (8, 9). In the UK, developing new approaches to the management of frailty has been facilitated by the adoption of the electronic Frailty Index (eFI) which identifies older people living with frailty based on their primary healthcare records (10). Older adults with frailty are much more likely to develop pain which impacts on their function compared to those without frailty (11).

The impact of pain in older adults with frailty is potentially modifiable. Older adults may particularly benefit from non-pharmacological strategies including physical, psychological, and social interventions, because changes in medication can have a disproportionate and negative impact on those with frailty (12). Whilst some older adults can self-manage their pain or be supported to do so by their general practice, many older adults may benefit from more specialised support. However, the high prevalence of pain in this population suggests that effective pain management remains a largely unmet need. Contributing factors may include older adults’ decisions not to seek treatment, inappropriate clinical decisions, or insufficient access to appropriate services (3). Pain services in the UK vary widely (4), spanning primary, secondary, and tertiary care, including NHS and non-NHS providers, and some with third-sector input. There is substantial regional variation.

The Pain in Older People with Frailty Study (POPPY) was a mixed-methods, co-design study which aimed to develop intervention content, implementation strategies, and professional guidance to support older adults manage their persistent pain (13). A participatory research approach was adopted throughout (14). This involves an active partnership between researchers and people affected by the research topic or involved in actions relating to the issues examined, in this case older adults and health care professionals (HCPs). The study consisted of four phases: in phase one, evidence of pain management programmes (PMP) and psychological therapies were reviewed (15). During phase two, the experiences of older adults living with frailty and pain were explored through qualitative interviews. In phase three, researchers conducted interviews with HCPs and commissioners across various pain service settings, locations, and provider types. The objective was to obtain a comprehensive understanding of the challenges and enablers associated with incorporating and managing older adults with frailty across various pain service types, and to identify the resources necessary to support them effectively. Phase four comprised co-design workshops with older adults and HCPs from a variety of service types across four locations to inform interventions that meet the needs of older adults living with frailty. This paper focuses on phase three, other phases of the POPPY study are reported elsewhere (13, 15).

## Aim

To capture the views of HCPs and commissioners relating to the barriers and facilitators to including and managing older adults with frailty in pain services, within different service contexts. Additionally, to gain insights from HCPs within generic community services to understand how they might support this population.

## Methods

### Design

In this phase we adopted a descriptive qualitative approach. This involved using in-depth semi-structured interviews with HCPs and commissioners in pain and community services across England, UK. The study is reported in accordance with the Consolidated criteria for Reporting Qualitative research (COREQ) guidelines (16) (see Supplementary Information 5 for completed checklist).

### Service identification and consent

To identify pain services, we drew on the research team’s expertise, reviewed previous pain service audits, conducted NHS website searches, and received support from the National Institute for Health and Care Research, Research Delivery Network. Pain services were purposively selected to represent primary, secondary, and tertiary care, non-NHS providers, and those with third-sector input. Additionally, two generic community services were identified that regularly included older adults with frailty and pain, to understand their approaches to supporting and managing this population.

Interested services were provided with detailed study information, followed by a Microsoft Teams call between the POPPY study co-lead (LB) and the service manager or their designate. After providing informed consent, the service manager/designate completed a questionnaire to supply background information about their service, treatment options, personnel, and a broad profile of their service populations.

### Service personal identification and consent

On behalf of the researchers, service managers purposively selected HCPs to ensure variation in roles and professional backgrounds and invited them to participate in interviews. Pain service managers were also asked to provide contact details of any commissioners who were involved with their service. POPPY researchers then sent interested HCPs and commissioners information sheets and consent forms. Participants provided informed consent by either returning a signed consent form or providing verbal consent which was audio-recorded prior to the interview. There was no contact between POPPY researchers and participants prior to the study. The aim was to interview 5–8 participants per service, accommodating fewer participants where service pressures limited availability. This ensured a range of personnel was included to reflect service type.

### Semi-structured interviews

Interviews were conducted by NK, NH and RM between February 2023 and November 2023, by telephone or videoconference (i.e., Microsoft Teams/Zoom). They took place at a time to suit the participant with no-one else present. No repeat interviews were undertaken. Interviews were audio-recorded with the participants’ permission and then transcribed verbatim. No fieldnotes were taken. Researchers were aware of the possibility that face-to-face interviews could elicit different responses to those conducted via telephone. However, most interviews were completed via videoconference, with telephone interviews only undertaken with HCPs unable to access the relevant technology. Because numbers of telephone interviews were small, and researchers detected no clear difference in responses between these two approaches it was decided that using data derived from both methods was acceptable. Topic guides for HCPs and Commissioners were informed by the POPPY study Programme Management Group (PMG), findings from earlier phases of the programme (13, 15) and adapted to reflect the different service types (see Additional files 1 and 2 for topic guides). During their development interview guides were piloted with the PMG which included HCPs currently working in pain services. with Additionally, prior to the interviews, researchers shared case studies with participants featuring older adults identified as frail (see Additional file 3 for case study example). Case studies were developed with the assistance of Patient and Public Involvement (PPI) group members, from interviews undertaken in a previous phase of the POPPY study with older adults aged seventy-five and over, and from PPI group members’ own experiences of living with pain (13). Case studies were referred to during interviews and served to prompt discussion and reflection on working with older adults with pain and frailty, recognising that not all pain services work extensively with this population. When introducing the aims of the interviews to participants, age parameters relating to the term “older adult” were deliberately undefined to avoid restricting the range of discussion and because people biologically age at different rates (17).

### Analysis

A thematic approach (18) to data analysis was used to explore the views of those interviewed. Data were managed without the use of data analysis software so researchers could maximise their familiarisation with the data and to facilitate the process of analysis (19). Firstly, three researchers (AW, NK, and NH) familiarised themselves with a subset of transcribed data comprising three transcripts. Researchers independently applied codes to the data. In keeping with the thematic approach, some of these codes were developed deductively, being based on topic guide questions and research objectives, whereas other codes emerged inductively from the data content. Following discussion and comparison of coded transcripts, consensus was reached on which codes best reflected the data and the first iteration of a coding framework was created. This initial coding framework was tested on a further subset of transcripts and refined through discussion before being applied to the remaining transcripts (See Additional file 4 for final coding framework). Once all transcripts had been coded, data were re-organised into themes, summarised, further refined and interpreted. Researchers (AW, NK and NH) met regularly to debate emerging themes, paying careful attention to negative cases. This iterative process continued until no further insights emerged from the data and saturation appeared to be achieved (20). To further enhance rigor, research methods and findings were regularly discussed with members of the PPI group and PMG, both of which met for the duration of the study. Transcripts were not shared with participants since they had the opportunity to feedback on the findings during the Phase 4 focus groups (to be reported elsewhere).

## Results

We approached fourteen services. One service declined due to capacity constraints. Two services initially agreed to participate but did not complete the process through to the interview stage. No specific reasons were provided, but we assume this was due to workload pressures. Eleven services successfully participated. The included services were in Yorkshire, Lancashire, Greater Manchester, Merseyside, East Sussex, and Devon. Nine of the services were pain services. Pain service settings included tertiary (specialist) hospital secondary care, community settings, and voluntary/third-sector components. Seven pain services were provided or partially provided by the NHS and two services were provided by a private organisation. All services were free at the point of access for patients. In addition, we included two non-pain services. One was a generic community service for older adults, providing care in the community. The other was an intermediate care, community-based service, responsible for the rehabilitation of patients registered with local Primary Care Network general practitioners (GPs). Both services regularly worked with older adults experiencing frailty and persistent pain.

Out of the 72 HCPs invited, twenty-two did not respond and six either declined to take part or did not complete their interview. A total of 44 interviews were completed with a range of commissioners and HCPs, e.g., physiotherapists, occupational therapists, nurses, psychological therapists, health and wellbeing coaches, GPs, a service manager, pain consultant, pharmacist, and a social prescriber (see Table 1). Interviews took between 45 and 75 minutes to complete. Findings from these interviews were used to inform workshop discussions in Phase 4 of the POPPY study (13). The workshops included many of the phase 3 interview participants alongside other pain service workers and contributed to guidance supporting the development of pain services for older adults with frailty.

**Table 1.** Participant professional roles.

| Professional role | Pain services |  |  | Generic | Totals |
| --- | --- | --- | --- | --- | --- |
|  | Tertiary | Secondary care | Community | Community services |  |
| Commissioner |  | 1 | 1 |  | 2 |
| Pharmacist | 1 |  |  |  | 1 |
| Physiotherapist | 3 | 11 | 2 | 2 | 18 |
| Occupational therapist | 1 |  | 1 |  | 2 |
| Nurse | 1 | 2 | 1 | 1 | 5 |
| Pain consultant |  | 1 | 1 |  | 2 |
| Psychological therapist |  | 3 | 2 |  | 5 |
| General practitioner |  |  | 2 | 1 | 3 |
| Social prescriber |  |  | 1 |  | 1 |
| Health / Wellbeing coach |  |  | 3 |  | 3 |
| Service manager |  |  | 1 | 1 | 2 |
| **Totals** | 6 | 18 | 15 | 5 | 44 |

## Findings

Two key themes emerged from the data: “*Characteristics shared by older adults with frailty shaping interactions with pain services*” and “*Factors likely to increase the effectiveness of pain services for frail older adults*”. Sub-themes relating to these two main themes were also identified (see Table 2).

**Table 2.**
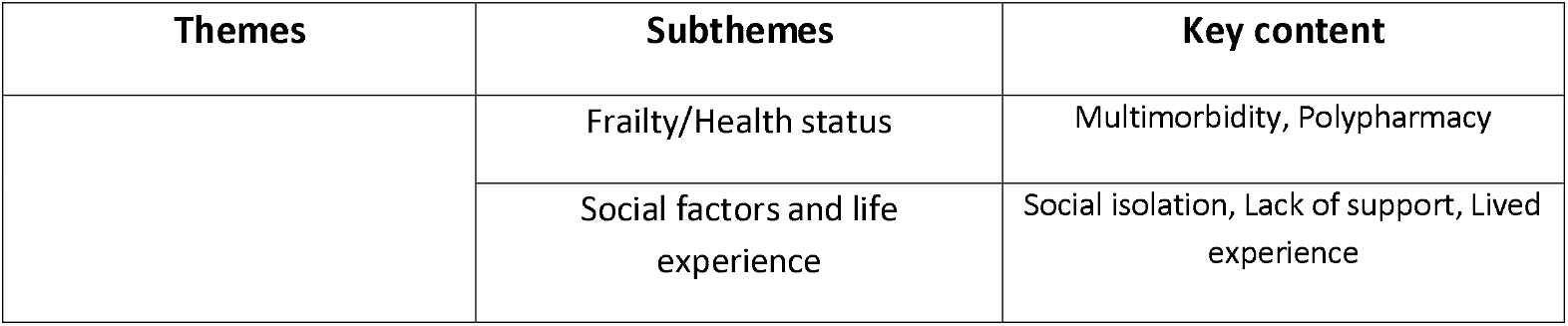
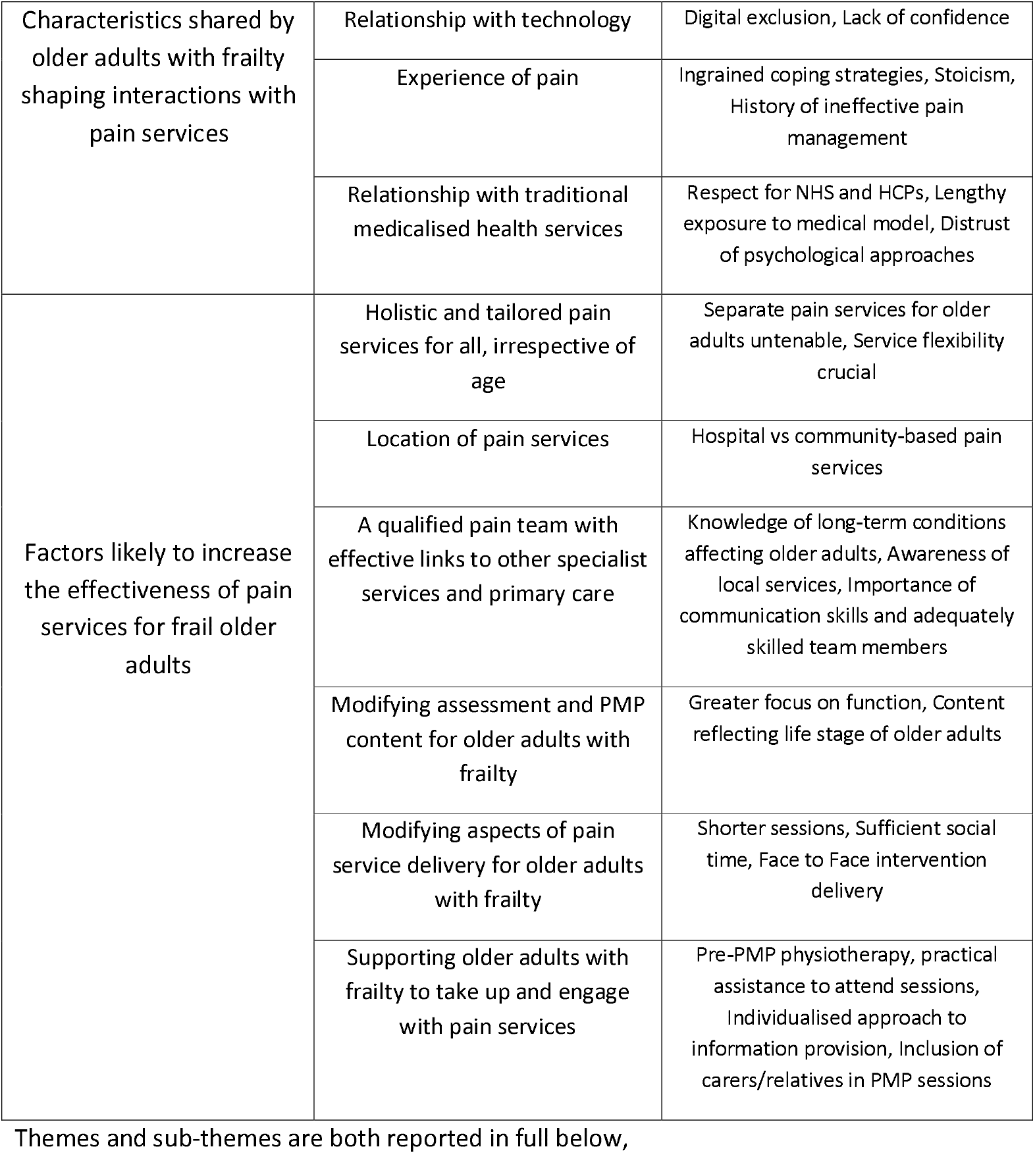
Summary of themes and sub-themes.

### Characteristics shared by older adults with frailty shaping interactions with pain services

Most HCPs recognised that older adults living with frailty and pain formed a cohort with some shared characteristics which influenced how pain services were delivered and how older adults engaged with them:

#### Frailty/Health status

The multiple and complex physical health issues experienced by many older adults with frailty presented a challenge for pain services. HCPs described how, what initially might appear to be a sensible course of action regarding medication changes, could impact other conditions and, therefore, had to be carefully considered. Similarly, the presence of polypharmacy could complicate adjustments to medication. Furthermore, pain services lacked the ability to respond to other health-related difficulties that older adults faced, and accessing frail older adults who were housebound or in care homes could be impossible,

> ***“Some people you never get to see because they are not able to come into clinic, so that can be quite difficult, you can’t even see how somebody walks or doesn’t walk so it’s not ideal”* (Pain Service occupational therapist)**

HCPs noted that older adults were likely to present with multimorbidity and this could make engagement with pain services difficult,

> ***“The type of older person that we get, and the frailty, the loss of that bounce-back ability when they get hit with something. I think that’s a big barrier. Or the co-morbidities flaring up or becoming generally unwell with something. That can set them back”* (Pain service physiotherapist)**

Uncontrolled pre-existing pain could be provoked through attending and participating in PMP activities, and a lack of mobility and continence issues could make participation in sessions problematic. Furthermore, cognitive impairment could lead to difficulties absorbing and retaining new information,

> ***“You’ve got the fact that older people can be generally frailer and are going to have more medical conditions and things like memory problems and may struggle with self-directed exercises”* (Pain service GP)**

#### Social factors and life experience

Participants recognised that although frail older adults’ lives could be intertwined with spouses or carers, many were socially isolated, lacked social support and were less autonomous than younger adults. Consequently, they thought older adults often lacked sufficient practical and emotional support to engage with pain services which could make the prospect of group sessions daunting at first. Conversely, participants believed that the social isolation experienced by many older adults could often provide the impetus to engage. Furthermore, they thought once older adults had accessed a PMP within a service, they had some advantages over younger patients: their wider experiences of life helped them to grasp concepts; they were less inhibited than younger patients and more likely to have the time to attend PMP as they no longer worked. Older adults were, therefore, often more likely to embrace group sessions than younger patients,

> ***“They’re (older adults) more comfortable accessing those type of things (pain groups) and, they bring all that stuff to the group. They’ve tried everything and they’re more open, they’ve accepted this is what’s happening”* (Pain service operations co-ordinator)**

Participants thought these factors also contributed to some older adults being more likely than younger patients to attend 6 months’ follow-up appointments.

#### Relationship with technology

With some exceptions, older adults were considered less accustomed to modern technology than younger people and, therefore, less confident using it to access pain services. Difficulties included accessing groups online and navigating information sent by email,

> ***“Being older doesn’t mean you haven’t got the ability to use technology, but it does sometimes mean that if you’re of a certain age and you haven’t used that technology before, it can be quite daunting, and you can be less willing to want to use that technology”* (Pain service clinical pain specialist)**

#### Experience of pain

Participants saw many frail older adults who had been living with pain for decades, during which time they had developed coping strategies that had become engrained, and which could help or hinder their interaction with pain services. For instance, they thought older adults could be more accepting of living with pain than younger adults and they felt sometimes a stoical view of their pain could work to the older adult’s advantage,

> ***“I don’t think the levels of distress and depression have been as high as maybe in the younger population, and that’s to do with life stage, and being a bit more accepting that pain is part of ageing”* (Pain service physiotherapist)**

Participants perceived that some older adults felt there was little point in seeking help at their age or that their acceptance of pain and lack of assertiveness in seeking help meant that by the time they eventually accessed pain services they were in great need,

> ***“If I had to rate who needed our input, older adults genuinely are often in a much more higher need group, not because of themselves, but because they kept quiet for so long before they’ve accessed our service”* (Pain service consultant)**

Participants reported that frail older adults’ long history of pain and associated treatment could result in them harbouring low expectations of pain services which could present a challenge to those delivering them,

> ***“Sometimes when older adults have lived with pain for so long, they sort of feel, well, I’ve tried everything, what’s talking about it going to do, sometimes it’s a bit more difficult to sort of get them on board with the holistic approach”* (Pain service social prescriber)**

#### Relationship with traditional medicalised health services

Participants perceived that many older adults highly valued the NHS and its workforce,

> ***“They say: “You’re the experts in your field”. They respect that; they’re kind of putting you on a pedestal”* (Pain service Psychological Therapist)**

Consequently, they tended to be respectful towards HCPs, were often eager for information and tended to follow advice,

> ***“They’re (older adults) keen to be here, they’re on time, they’re ready to go. You never get an older adult getting their phone out in the middle of the session. They’re so respectful”* (Pain service physiotherapist)**

This respectful attitude made older adults popular with HCPs delivering pain services. However, participants believed that a long exposure to the medical model of pain management could shape older adults’ expectations and lead to a reluctance on the part of some older adults to take control. They tended to defer to medical “experts” and preferred a medicalised approach to pain management in which clinicians provided solutions in the form of a physical intervention or change in medication. Participants felt this led to a mismatch between patient expectations and the non-curative ethos of pain services which aimed to facilitate self-care and self-management. Participants believed some older adults were resistant to shifting from the expectation of a cure,

> ***“For some patients it’s quite a shift, if you’ve been going to lots of consultations and it’s been about trying to reduce your pain and then you come to a clinic and somebody’s saying, ‘Actually, we’re not going to reduce the pain, we’re going to work with you to manage the pain and be able to live in the presence of pain’”* (Pain service occupational therapist)**

Participants thought that older adults struggled at times with understanding abstract concepts relating to pain,

> ***“I think for a lot of older adults, grasping the modern concepts around pain can be harder because they’ve had longer to have this misunderstanding around pain to be very much ingrained in them by health professionals, the media, etc. So, I think it can be hard to undo some of the damaging effects of that”* (Pain service physiotherapist)**

Participants perceived that older adults could also be a “harder nut to crack” and less malleable than younger patients because they had been told for a long time that their pain was an untreatable physical phenomenon that they had to put up with,

> ***“They think pain is inevitable because of what they have been told by clinicians; you have to expect to be less mobile when you’re older, meds don’t work as well for older adults, you’re bound to get more side effects”* (Pain service consultant)**

Participants thought decades of exposure to the biomedical model of care and unfamiliarity with concepts such as “Mindfulness” could lead to older adults holding a negative attitude towards psychological approaches in contrast to younger adults, amongst whom the concept of psychology has been normalised,

> ***“I think the team have a strong sense about psychological approaches feeling more alien to the older generation. Older people may be less open to biopsychosocial approaches, or it might just feel so far away from what they thought was going to happen if they’ve had a very medical journey with their pain”* (Pain service clinical psychologist)**

Participants felt that older adults tended to associate psychology with mental illness and the stigma they attached to this, although participants did admit that outcomes from one-to-one psychology services were as good as with younger adults. A participant noted that some clinicians excluded older adults from psychological interventions, assuming these management methods were unfamiliar to them and be unlikely to succeed.

### Factors likely to increase the effectiveness of pain services for frail older adults

#### Holistic and tailored pain services for all, irrespective of age

Some participants, including those from non-pain services, thought a separate pain service would benefit older adults with frailty, arguing that it would be more responsive to their specific needs. Several cited frail older adults’ difficulties accessing existing services,

> ***“A lot of these frail people can’t get to the class I think when somebody is older and they’ve got all these comorbidities, sometimes they just put up with pain as that’s the least of their worries because, they’re struggling to breathe or they’re struggling to walk”* (Non-pain service physiotherapist)**

Most participants, however, were not in favour of a separate service either because pain services already accounted for the needs of older adults and were well attended, or there were insufficient frail older adults in their locality who would potentially benefit from a separate service to justify creating one,

> ***“It’s a numbers game, isn’t it, that’s what it comes down to in the NHS. If you’ve got enough people coming through that would support a specialist service, then maybe you could do that. For me, we don’t have the numbers”* (Pain service physiotherapist)**

Some participants suggested that pain services should be flexible enough to focus on patients’ unique circumstances and meet the needs of all adults irrespective of age and frailty state,

> ***“I wouldn’t go in and think, right you’re over 75 so I must use a different approach for you. I would adapt my approach depending upon what they shared with me they wanted to work with in the same way that I would with someone who’s under 75”* (Pain service psychological therapist)**

For instance, having a tailored approach could allow patients to be allocated six, twelve or eighteen hours of a PMP depending on their previous experience with pain and current presentation, regardless of age. Some suggested that the negative connotations of a service specifically for older adults could put potential patients off and risked marginalising those older adults using it. One HCP suggested that a separate service would be less effective,

> ***“I think with a very specialist service you can end up being very prescriptive in what you do, because you lose sight of the bigger picture”* (Pain service physiotherapist)**

Several participants suggested that in creating a separate service for older adults, the benefits from having patients of mixed ages attending groups would be lost. Some thought having older adults in groups alongside younger adults enhanced its effectiveness because older members often adopted a parental role, imparting knowledge and advice learned through experience to younger members,

> ***“I think it’s always really nice when we were doing the face-to-face groups, that the young and the old mixed together, they shared stories, they supported each other, they both had a purpose and a role there”* (Pain service wellbeing practitioner)**

Participants thought there were practical problems around establishing and running a separate pain service. Recruiting sufficient staff would be problematic and risk diluting existing services,

> ***“I feel a little bit uncomfortable about a dedicated service. I think you’d struggle to recruit into a field of medicine that’s already difficult to recruit to if you made it more specialised, because you’re just narrowing the pool of people with an interest”* (Pain service physiotherapist)**

They also warned that a small, specialised service would be at risk from a lack of resources and prone to absences of key staff. Identifying age-related inclusion criteria for a separate group could also lead to problems,

> ***“Do you really want to marginalise people into a particular service by age? We see ninety-year-old people that are fitter than some twenty-five-year-olds. So would they be better served in a cohort of people that form their peer group in terms of age?”* (Pain service physiotherapist)**

#### Location of pain services

Some participants working in secondary care identified potential advantages associated with hospital-based pain services, particularly for managing severe acute pain. They thought it useful to have the pain team located at the same site as other relevant medical specialities (e.g., orthopaedics, radiologists, anaesthetists, and in-patient physiotherapy) and facilities such as hydrotherapy pools. Most participants, however, were strongly in favour of locating services for those with chronic pain in the community, because this fitted with the prevalent shift towards a biopsychosocial model of pain management,

> ***“We try and stay away from medical settings. For the pain management programmes, we try and keep them in the community. We’re trying to instil some self-efficacy rather than bringing them into a medical environment where most people will be under the preconception that something passive will be done to them for the pain”* (Pain service physiotherapist)**

One commissioner proposed a three-tiered approach to pain services, comprising education and signposting in the basic tier, one-to-one/group interventions in the second tier (both delivered in the community), and specialised services such as psychological services/medical interventions in the third tier (delivered in hospital settings),

> ***“You have a lower level (Tier 1) which is a catch all. That service is there to educate, signpost on and provide materials. The Tier 2 is the bit where you get more involved in groups or the one-to-one stuff and is for people that feel they can’t manage their pain themselves. That is probably the bit that most people will get most benefit from. Then the Tier 3 bit is for the people who identify of having a psychologically based issue that impedes their ability to manage their pain”* (Commissioner)**

Furthermore, commissioners suggested that building effective and accessible community-based pain services which met the needs of most patients experiencing chronic pain would allow hospital-based services to focus on those presenting with the severest problems,

> ***“I want a service that’s effective and delivers evidence-based care and that’s seeing the right cohort of patients and that’s why we need this community service because actually, most patients don’t need to go anywhere near secondary care, and then hospital services can focus on the really desperate patients that do need to be in secondary care”* (Commissioner)**

Some HCPs suggested that GP surgeries were better placed than hospital-based pain services when modifications to medications were indicated and patients needed support in adapting to these changes,

***“Medication changes should be happening more locally in GP practices rather than patients having to wait and come to see a secondary specialist miles away from where they live. They should be undertaken by someone who can connect them with stuff locally that might have an impact on their day-to-day living which all goes towards living better with the pain”* (Pain service pharmacist)**

Accessing hospital services were considered challenging for frail older adults, with participants citing the stress associated with navigating complex buildings, accessing parking and the discomfort of lengthy journeys,

> ***“Mobility is a huge issue for people and coming to a busy hospital like this where parking and everything is an issue, that adds to the stress”* (Pain service psychological therapist)**

They also thought pain services should be local to patients, and have local facilities including gyms, swimming pools and social groups for which social prescribing could be made more accessible,

> ***“A while back, our service was commissioned to develop a community pain service and we were able to deliver that from a community leisure centre. They had a gym there and other facilities that people might utilise in the future, and I think there are some benefits from that”* (Pain service occupational therapist)**.

#### A qualified pain team with effective links to other specialist services and primary care

Participants thought that equipping the workforce with relevant skills and knowledge was vital. Awareness of the long-term conditions likely to affect frail older adults was crucial due to the likelihood of multimorbidity,

> ***“I think we could do with more training from people with expertise in the older population on how we should be adjusting our assessments and our support. Training on working with people with dementia and the wide range of potential co-morbidities would also be helpful. Perhaps some of some of our non-medical members of the team like me and my psychology colleagues could do with more understanding of those medical aspects and how they might impact on someone’s pain” (Pain service psychologist)***

Also considered important was an awareness of local services and effective signposting,

> ***“It would be useful to have a better understanding of community services and charities within our organisation, and a better index of available services”* (Pain service clinical pain specialist)**

Having a suitable professional mix including social prescribers and psychologists, plus key specialist input from frailty leads and link nurses, was thought to be important. Participants from non-pain services were in favour of including GPs in pain teams. Employing staff with the skills and personalities to interact effectively with frail older adults who may face communication difficulties was also considered crucial,

> ***“There are certain staff members that maybe don’t work as well with older people, and particularly people who are frail. They need somebody that is calm, patient, and that has a rapport with them”* (Pain service wellbeing practitioner)**

Similarly, participants believed it was important to employ staff with languages relevant to the population served whenever possible. Several suggested that effective teamwork was dependent on having dedicated time to discuss caseloads with multidisciplinary teams including GPs and sufficient capacity to complete post-discharge reviews.

Citing the complexity of many frail older adults’ presentation, participants from all service types identified the importance of effective working relations with GPs, community services such as falls teams, social services and other medical specialities,

> ***“If it’s looking at any sort of invasive treatment, we may have to bring the pain consultant on board for things like complex case reviews because often, there isn’t a straight-forward answer or treatment for older people”* (Pain service physiotherapist)**

This might involve the development of integrated teams incorporating representatives from health, social services, and community/voluntary sectors. Participants thought that effective communication with GPs via electronic communication and education to ensure appropriate referrals was important. They also acknowledged that a responsive and flexible local voluntary sector helped when planning actions with older adults. HCPs from non-pain services reported they would like access to in-reach components of pain services to educate and support them in the management of people with chronic pain as they lacked confidence and skills with applying psychological approaches. In contrast, HCPs from some pain services wanted more training on frailty and the management of multimorbidity.

#### Modifying assessment and PMP content for older adults with frailty

Some participants reported adjusting their approach when assessing frail older adults as opposed to younger adults, with a greater focus on function and mobility,

> ***“If I get a frail older adult referred and they’re interested in the pain management programme, I’ll bring them in for an assessment to check their mobility, what their transfers are like, what they are doing”* (Pain service physiotherapist)**

Participants thought intervention content should reflect the likelihood of multiple underlying medical conditions and a different stage of life to younger adults,

> ***“In our “Understanding your body” session, when we talk about different structures in the body and how they work, there’s more emphasis on explaining joint related pain because that tends to be more prevalent in our older adults’ groups. Also, we do more balance and movement exercises rather than Pilates”* (Pain service occupational therapist)**

They believed that information was more likely to be understood by older adults when simplified. Similarly, PMP content should be geared specifically towards older adults, for example, retirement issues rather than a return to work,

> ***“We won’t do “Work and Employment” in the older adult programme but we do do sessions around staying productive and leisure activities that we link with social prescribing”* (Pain service occupational therapist)**

Information about bone density in older age was thought to be particularly relevant to older adults,

> ***“We talk about bone density and what preserves it and what doesn’t. Weight bearing is very necessary, your body won’t stimulate osteoblasts to ossify, calcify and build strong bones unless you weight bear”* (Pain service physio)**

Participants thought that pain education sessions should include expectations of pain in the older body, and discussions of medical conditions such as osteoarthritis should be specific to older adults and avoid terms such as “wear and tear” and “degeneration”,

> ***“What we are trying to do is explain every single pathology in a way that enables that older patient, not frightens them, not disables them. So, osteoarthritis would never be described as “wear and tear” or “degenerative” because although those are terms that people use all the time, they’re not that accurate and they do actually promote disability”* (Pain service physio)**

Several participants commented on the importance of groups and peer support and recommended introducing an older adult who had successfully engaged with pain services to PMP groups to describe the benefits they had experienced,

> **“*We have who we call our “poster boy”. He’s an older gentleman, probably mid-60s, and he’s a perfect example of a person that previously wasn’t really that bothered, just wanted the medicines, but then having received the service his life has completely changed around. He will walk in a room, tell them what has happened to him, and they will probably go and sign up to everything that he suggests*” (Pain service manager)**

Participants at one location described a separate PMP for over 65-year-olds that had been created in response to older adults’ difficulties with mobility and a recognition that the content of the standard PMP did not resonate with this age group. They did acknowledge, however, that filling these groups could take considerable time due to the small number of older adults who would benefit from them being referred to their pain service,

> **“*I’d say waiting for sufficient referrals before launching courses for over 65 groups is a little downside because although you get a programme that’s more appropriate with content very tailored to their needs, sometimes the wait is longer*” (Pain service physiotherapist)**

#### Modifying aspects of pain service delivery for older adults with frailty

Participants believed interventions should be modified to reflect the reduced tolerance for physical activity that many frail older adults with frailty experienced. They felt it was important to reduce session content and include more breaks when interacting with older adults. They also noted that more time was needed with this age group as older adults were often socially isolated and appreciated the opportunity to talk about their pain and reflect on it with others,

> **“*I always ask them to tell me about their pain journey because often, they haven’t been given the time to explain what’s brought them to this point, and I particularly find that very hard for the older generation. They want to offload, and that purpose of offloading and being heard and then sounding it back to them, gets their engagement”* (Pain service wellbeing practitioner)**

While some participants recognised that frail older adults often liked the convenience of a telephone assessment where they could sit at home and avoid the cost and burden of travel, others were strongly in favour of face-to-face interaction with older adults, when possible, particularly when working with people who had sensory loss and when talking about abstract concepts. Participants from several services reported that their delivery had become telephone-based during the COVID-19 pandemic and were now reinstating face-to-face sessions. They thought that older adults preferred this approach too,

> **“*I think most older people would prefer face-to-face; I think it’s a little bit more old school, but people generally like to do business face-to-face. And I think particularly with people who may have hearing issues, or any form of communicative difficulty it’s easier negotiated if you’re face-to-face*” (Pain service physiotherapist)**

Participants were also in favour of offering frail older adults the option of community-based appointments which they thought may suit patients with communication/sensory difficulties and home visits, particularly for older adults who were housebound or residing in assisted living facilities.

Participants across all service types recognised that groups conferred social benefits for older adults such as peer support, particularly for those experiencing social isolation,

> **“*PMP seems to help the older adults because they’ve had these pains for years, for twenty years, thirty years and they’re just living with it, but no one’s really told them how to manage it, so they’ve never really had the support that they need. So, when they come in and they share their experience they look at each other and are like ‘Oh, I’m not the only one in pain’*” (Pain service physiotherapist)**

#### Supporting older adults with frailty to take up and engage with pain services

Participants recognised that frail older adults may need extra input to prepare them for attending a PMP. This could include being seen individually for reassurance or community physiotherapy visits to ensure they had sufficient skills and resilience. Most commented that older adults were often in need of practical assistance with transport to attend sessions. Ideally, this should be through the provision of taxis due to the difficulties faced by many older adults with tolerating hospital transport,

> ***“I have had many patients saying: “I’d like to come to clinic, but you know what happens with the transport’ they go all around the houses and they take so long to get home I couldn’t hold on for a wee that long”“ (Pain service clinical pain specialist)***

Participants advocated posting out hard copies of leaflets and literature to older adults who were struggling with technology and consequently could not access electronic versions,

> ***“The other day I was talking to a lady about mindfulness. I couldn’t email her the links so she could listen to the exercises because she didn’t have the internet. Fortunately, we were able to post out a handbook to her and she said, “I can read the three-minute breathing exercise and understand it but I’m not relaxing because I’m reading it”. So, I printed her a CD and she’s found it so beneficial she’s using it daily” (Pain service wellbeing practitioner)***

Participants from one service described routinely telephoning patients two weeks after despatching information through the post to see if they had received and understood it and to ask whether they had any further questions. Others reported providing older adults who lacked access to technology with recorded instructions for exercises.

Most participants were in favour of including carers or relatives in one-to-one interactions with older adults when emotional support was needed or there was a risk of the older adult forgetting information,

> **“*We try to involve family for support and advocacy. Particularly when issues are quite complicated or there’s a feeling that understanding the information is a challenge. If people are clearly not understanding what we’re saying, having someone to advocate, or a second pair of ears or somebody who can go away and talk to them is really useful*” (Pain service physiotherapist)**

Some also included carers and relatives in PMP sessions on rare occasions for the same reasons, although they did admit this could potentially lead to issues around confidentiality.

## Discussion

The overarching aim of the POPPY study was to develop guidance relating to pain service design, intervention content, and implementation strategies to enable older adults with frailty to manage their pain more effectively. In phase 3 of POPPY, we set out to understand the barriers and facilitators influencing how older adults with frailty engage in pain services from the perspective of HCPs and commissioners and explore their views on how pain services for this population can be improved. Through the emergence of two key themes: “*Characteristics shared by older adults with frailty shaping interactions with pain services” and “Factors likely to increase the effectiveness of pain services for frail older adults*” we have fulfilled our original study aims and contributed to current knowledge in both these areas.

### Barriers and facilitators to engagement in pain services

While recognising that older adults with frailty did not constitute a homogenous group, study participants identified several shared characteristics among older adults that differed from younger adults and which shaped their access to and interaction with pain services in the form of barriers and facilitators. Certain common characteristics that emerged such as stoicism and a desire for social contact exerted a positive influence and have been noted elsewhere (21). This aligns with other published research recognising that older adults with pain have specific needs and that managing pain in this population is different to younger people (5). Despite differences between older adults and younger age groups, there are few published guidelines that relate specifically to older adults (22). For example, current chronic pain guidance for adults recognises the consideration of a subgroup of younger adults (16-25 years), but not older adults (23). However, clinicians are advised to consider unique challenges to pain management due to characteristics including both children and older adults (≥65 years) (24, 25).

Our findings suggest that frail older adults may expect medicalised approaches to their pain management (26) and could be wary of psychological interventions, either due to lack of familiarity with them or because they associate psychology with the management of mental illness rather than chronic pain. Furthermore, older adults’ lack of contact with psychological approaches may be exacerbated by HCPs’ assumptions about older adults’ lack of willingness for psychologically informed therapies. Consequently, this limits the numbers referred to mental health services. Other studies have found that older adults’ willingness to engage in non-pharmacological pain management strategies was aligned to their awareness of these strategies, their appeal and accessibility (27). Part of the issue may be a lack of application of biopsychosocial approaches to managing older adults’ pain among clinicians (27). Issues around knowledge translation and implementation into clinical practice could be a factor when moving from long established biomedical models towards biopsychosocial pain management for older adults with frailty. Recommendations for knowledge translation projects for pain management for older adults include pain management education for older adults, information on the safe use of pharmacology, and self-management strategies (28). However, a review of knowledge translation programmes aimed at clinicians working with older adults found that while such programmes raised awareness and knowledge among clinicians, this did not always change clinical practice (29).

We found that older adults with frailty may experience difficulties with online contact with pain services such as engaging with electronic information. Recommendations have previously been made to improve usability for older adults through considering their cognitive and physical abilities in the design of electronic resources, for example, the navigation pages, text size and buttons (30).

### Improving pain services for older adults with frailty

Participants suggested that building pain service clinicians’ confidence in delivering psychologically informed therapies was important. This is vital for physiotherapists, who because of their undergraduate education, may tend to adhere to a biomedical perspective when assessing and treating patients with chronic pain (31). Participants’ view that increasing the knowledge of pain management in non-pain services was important, is mirrored by recent published recommendations (32). Additionally, it is crucial that pain team staff possess adequate knowledge and skills of the clinical impact of older age and frailty to inform their treatment planning. Our findings suggest that there is a need for effective interdisciplinary working between all HCPs, including GPs, community services and specialist services who all have a vital role to play in assessing older adults with frailty. This would ensure older adults receive a consistent message about the possible aims of pain management, while also being referred to the appropriate and available services in their locality.

The view of the participants, that multidisciplinary team (MDT) led approaches were needed, is generally accepted (5) and echoes current standards and recommendations (32) (CSPMS 2021). Although it is also recognised that a significant barrier to achieving a quorate MDT is the difficulty in accessing sufficient physiotherapists and psychologists (CSPMS 2021). This is particularly significant as psychologists and physiotherapists are often key personnel in the delivery of chronic pain interventions (31, 33)

Rather than establishing specific services for older adults with frailty and pain, participants favoured embedding the care of this population within existing pain services, while ensuring that this service was patient-centred, flexible in delivery, tailored and age-appropriate. The importance of tailoring pain service delivery to the individual, even in group interventions, is well accepted (15, 23, 33, 34).

Participants were in favour of optimising the location of pain services to minimise travel and offering practical assistance with transport for older patients with frailty. Participants also advised on the importance of social prescription to accessible community and voluntary services. This aligns with previous findings that transportation, cost and accessibility should be considered when planning pain management programmes for older adults (15, 28).

While recognising the need for specialist secondary and tertiary pain services to be hospital based, participants thought that community-based pain services would be less medicalised and, therefore, more likely to promote self-management. Others have noted similar benefits and have also suggested a lower cost compared with hospital-based services (35).

### Strengths and limitations

Our engagement with a variety of clinical service types and locations across England, and the completion of interviews with a wide range of clinicians and commissioners, represents a strength of our study. Consequently, we gained considerable insight into clinical perspectives of the barriers and facilitators to older people with frailty accessing pain services and improving pain services for older adults. For this study, we purposefully did not define ‘older adults’ by a specific age. Rather, we used an inclusive approach to considering the impact of frailty and pain since this may vary across individuals, beyond age categories. The qualitative findings will be used to develop recommendations for a pain service model for this population.

We relied on service managers to identify potential participants from their locality and invite them to be interviewed. Inevitably, this approach may have introduced a degree of participant selection bias. The interviews were undertaken by researchers who did not have a clinical background, and the data were interpreted by the wider research team with mixed research and clinical backgrounds (including musculoskeletal/pain specialism physiotherapy and general practice). There will be some researcher influence on the interpretation of the findings based on their interest and their experience of the barriers in the management of chronic pain for older adults with frailty from their backgrounds in primary and secondary care. We did not collect demographic data regarding the participants and so we cannot report on their ages/ethnic backgrounds/clinical experience. However, characteristics of one of the services delivered by our participants were adapted to reflect the multi-cultural nature of the community served, e.g., by recruiting staff who were multi-lingual and of similar faith to the local population and locating community-led support groups in a variety of faith centres. Future study can explore the barriers and facilitators to accessing pain services among more diverse groups of older adults living with frailty.

## Conclusion

The way older adults with frailty interact with pain services is shaped by a range of factors that require careful consideration if their specific needs are to be met. Within existing pain services there is a call to upskill HCPs with the knowledge and skills to meet the needs of this complex and vulnerable group. While a separate service for older adults with frailty may not be required, modification to existing services may be needed, alongside the provision of tailored treatment and the practical support necessary for older adults to engage in pain services.

## Supporting information

Supplementary File1 Topic Guide HCPs

Supplementary File 2 Topic Guide HCPs & Commissioners

Supplementary File 3 Case Study

Supplementary File4 Final Coding Framework

COREQ Checklist

## Data Availability

All data produced in the present study are available upon reasonable request to the authors

## Abbreviations

GP: General Practitioner
HCP: Health Care Professional
NHS: National Health Service
PMP: Pain Management Programme
POPPY: Pain in Older People with Frailty Study

## Supplementary Information

Additional file 1: Interview Topic Guide HCP (.pdf)

Additional file 2: Interview Topic Guide Commissioners (.pdf)

Additional file 3: Case Study (.pdf)

Additional file 4: Final Coding Framework (.pdf)

Additional file 5: Completed COREQ checklist (.pdf)

## Declarations

## Acknowledgements

The POPPY Study Programme Management Group: Prof Amanda Williams, Prof Patricia Schofield, Dr Asim Suleman and Dr Deborah Antcliff.

The participating services.

The National Institute for Health Research, Research delivery Networks (known formally as the Clinical Research Network)

The POPPY study Patient & Public Involvement (PPI) group

## Authors contributions

NK (Senior Research Fellow, female), NH (Research Fellow, female) and RM (Research Fellow, female) conducted the interviews. They have substantial experience of qualitative data collection. Data analysis was undertaken by AW (Research Fellow, male), NK and NH. NK and AW have substantial experience of qualitative data analysis. The manuscript was prepared by AW and DA (Advanced Physiotherapy Practitioner/Clinical Research Fellow, female) with additional input from AS (General Practitioner with an extended role in Pain Management, male). All other authors were involved in the conception and development of the POPPY trial. AW, DA, NK, RM, LB (POPPY Co-Lead), and AF (POPPY Co-Lead) have PhDs. All authors read and approved the final manuscript. The views expressed are those of the author(s) and not necessarily those of the NHS, National Institute for Health and Care Research or the Department of Health and Social Care.

## Availability of data and materials

All data generated or analysed during this study are available from the corresponding author on reasonable request.

## Ethics approval and consent to participate

The study protocol was approved by Leeds-East Research Ethics Committee on 28 April 2022 (22/YH/0080).

## Consent for publication

Not applicable.

## References

1. Abdulla A, Adams N, Bone M, Elliott AM, Gaffin J, Jones D, et al. Guidance on the management of pain in older people. Age and ageing. 2013;42:i1–57.

2. Van Hecke O, Torrance N, Smith B. Chronic pain epidemiology and its clinical relevance. British journal of anaesthesia. 2013;111(1):13–8.

3. Reid MC, Eccleston C, Pillemer K. Management of chronic pain in older adults. Bmj. 2015;350.

4. Kailainathan P, Humble S, Dawson H, Cameron F, Gokani S, Lidder G. A national survey of pain clinics within the United Kingdom and Ireland focusing on the multidisciplinary team and the incorporation of the extended nursing role. British Journal of Pain. 2018;12(1):47–57.

5. Ong T, Thiam CN. Special consideration for pain management in the older person. Clinical Medicine. 2022;22(4):295.

6. Brown L, Young J, Clegg A, Heaven A. Pain in older people with frailty. Reviews in Clinical Gerontology. 2015;25(3):159–71.

7. O’Caoimh R, Sezgin D, O’Donovan MR, Molloy DW, Clegg A, Rockwood K, et al. Prevalence of frailty in 62 countries across the world: a systematic review and meta-analysis of population-level studies. Age and ageing. 2021;50(1):96–104.

8. Harrison JK, Clegg A, Conroy SP, Young J. Managing frailty as a long-term condition. Age and ageing.44(5):732–5.

9. Department of Health. NHS Long term plan 2019 London: Department of Health; 2019 [Available from: https://www.longtermplan.nhs.uk/

10. Clegg A, Bates C, Young J, Ryan R, Nichols L, Ann Teale E, et al. Development and validation of an electronic frailty index using routine primary care electronic health record data. Age and ageing. 2016;45(3):353–60.

11. Brown L, Young J, Teale E, Santorelli G, Clegg A. A Cross-Sectional Study of the Impact of Pain in Older People with Frailty: Findings from the Community Ageing Research 75+(CARE75+) Study. Advances in Geriatric Medicine and Research. 2019.

12. Clegg A, Young J, Iliffe S, Rikkert MO, Rockwood K. Frailty in elderly people. The lancet. 2013;381(9868):752–62.

13. Brown L, Mossabir R, Harrison N, Lam N, Grice A, Clegg A, et al. Developing the evidence and associated service models to support older adults living with frailty to manage their pain and to reduce its impact on their lives: protocol for a mixed-method, co-design study (The POPPY Study). BMJ open. 2023;13(6):e074785.

14. Vaughn L, Jacquez F. Participatory Research Methods-Choice Points in the Research Process. Journal of Participatory Research Methods 2020; 1 (1)

15. Lam N, Green J, Hallas S, Forster A, Crocker TF, Andre D, et al. Mapping review of pain management programmes and psychological therapies for community-dwelling older people living with pain. European Geriatric Medicine. 2023;1–13.

16. Tong A, Sainsbury P, Craig J. Consolidated criteria for reporting qualitative research (COREQ): a 32 -item checklist for interviews and focus groups. International Journal for Quality in Health Care. 2007; 19 (6):349–357

17. NHS England. Improving care for older people [Available from: https://www.england.nhs.uk/ourwork/clinical-policy/older-people/improving-care-for-older-people/.

18. Braun V, Clarke V. Using thematic analysis in psychology. Qualitative research in psychology. 2006;3(2):77–101.

19. John WS, Johnson P. The pros and cons of data analysis software for qualitative research. Journal of nursing scholarship. 2000;32(4):393–7.

20. Naeem M, Ozuem W, Howell K, Ranfagni S. Demystification and actualisation of data saturation in qualitative research through thematic analysis. International Journal of Qualitative Methods. 2024;23:16094069241229777.

21. Washburn AM, Williams S. Becoming and being an older adult: A mixed methods study of the lived experience of aging. Journal of Aging Studies. 2020;54:100871.

22. Schofield P, Dunham M, Martin D, Bellamy G, Francis S-A, Sookhoo D, et al. Evidence-based clinical practice guidelines on the management of pain in older people–a summary report. British Journal of Pain. 2022;16(1):6–13.

23. (NICE) NIfCE. Chronic pain (primary and secondary) in over 16s: assessment of all chronic pain and management of chronic primary pain. 2021.

24. Cohen SP, Vase L, Hooten WM. Chronic pain: an update on burden, best practices, and new advances. The Lancet. 2021;397(10289):2082–97.

25. Cohen SP, Vase L, Hooten WM. Chronic pain: an update on burden, best practices, and new advances. Lancet. 2021;397(10289):2082–97.

26. Yarycky L, Castillo LI, Gagnon MM, Hadjistavropoulos T. Initiatives Targeting Patients: A Systematic Review of Knowledge Translation Pain Assessment and Management Studies Focusing on Older Adults. The Clinical Journal of Pain. 2024;40(4):243–52.

27. Garrett SB, Nicosia F, Thompson N, Miaskowski C, Ritchie CS. Barriers and facilitators to older adults’ use of nonpharmacologic approaches for chronic pain: a person-focused model. Pain. 2021;162(11):2769–79.

28. Yarycky L, Castillo LIR, Gagnon MM, Hadjistavropoulos T. Initiatives Targeting Patients: A Systematic Review of Knowledge Translation Pain Assessment and Management Studies Focusing on Older Adults. Clin J Pain. 2024;40(4):243–52.

29. Yarycky L, Castillo LIR, Gagnon MM, Hadjistavropoulos T. Initiatives Targeting Health Care Professionals: A Systematic Review of Knowledge Translation Pain Assessment and Management Studies Focusing on Older Adults. Clin J Pain. 2024;40(4):230–42.

30. Wildenbos GA, Jaspers MWM, Schijven MP, Dusseljee-Peute LW. Mobile health for older adult patients: Using an aging barriers framework to classify usability problems. Int J Med Inform. 2019;124:68–77.

31. van Dijk H, Köke AJ, Elbers S, Mollema J, Smeets RJ, Wittink H. Physiotherapists using the biopsychosocial model for chronic pain: Barriers and facilitators—A scoping review. International journal of environmental research and public health. 2023;20(2):1634.

32. Wildenbos GA, Jaspers MW, Schijven MP, Dusseljee-Peute L. Mobile health for older adult patients: Using an aging barriers framework to classify usability problems. International journal of medical informatics. 2019;124:68–77.

33. Joypaul S, Kelly F, McMillan SS, King MA. Multi-disciplinary interventions for chronic pain involving education: A systematic review. PloS one. 2019;14(10):e0223306.

34. Medicine FoP. Core Standards for Pain, Management Services in the UK second Edition 2021 [Available from: https://www.britishpainsociety.org/static/uploads/resources/files/FPM-Core-Standards-2021.pdf.

35. Simm R, Barker C. Five years of a community pain service solution-focused pain management programme: extended data and reflections. British Journal of Pain. 2018;12(2):113–21.

