## Supplementary File1 Topic Guide HCPs for "Maximising pain services for frail older adults, the views of healthcare professionals and commissioners: findings from the Pain in Older People with Frailty (POPPY) study"

**POPPY study**
**Topic guide for interviews with healthcare professionals**

**1. HCPs' experience of pain management services for older people**
1.1 Describe your experience of working in/referring to pain management services for older people, those with comorbidities and with frailty
1.2. What pain services are available in your area (i.e. within tertiary centres, secondary care, primary care)?
1.4 How effective do you think current pain services are for older people?
1.5 How effective do you think pain management programmes, or psychological therapies are for older people?

**2. HCPs' perceived barriers and facilitators for older people accessing pain management**
2.1 What factors may currently prevent older people, those with comorbidities and with frailty being referred to pain services?
2.2 What factors may facilitate older people's access/ referral to pain services?
2.3 What factors may prevent older people from attending pain services?
2.4 What factors may facilitate older people's attendance in pain services?

**3. Providing pain services for older people with frailty**
3.1 What factors need to be considered in developing services for older people with comorbidities and with frailty?
3.2 What would be the key components of pain services for older people, those with comorbidities and with frailty?
3.3 What would be the key components of a pain management programme for older people, those with comorbidities and with frailty?
3.4 What might be the similarities and differences between pain services for older people and pain services for younger people/without frailty?

**4. Tailoring to older people, those with co-morbidities and frailty**
4.1. What information would commissioners need to know when developing/funding pain services for older people, those with co-morbidities and frailty?
4.2. What extra training would HCPs need?
4.3. What other services need to be involved/linked in and how (e.g. medical specialities)?

**5. Improving accessibility**
4.3. What additional resources are needed to improve accessibility (e.g. written document font/braille, alternative formats-audible, other languages?)
4.4. What services in the community could link with NHS services and how can they support accessibility?

**6. Vignettes**
6.1. After reading the vignettes, how would you manage Patient X? (explain)
6.2. How would you manage Patient Y? (explain)

**7. General comments**
7.1. Do you have any additional comments to make?
