## Supplementary File 2 Topic Guide HCPs & Commissioners for "Maximising pain services for frail older adults, the views of healthcare professionals and commissioners: findings from the Pain in Older People with Frailty (POPPY) study"

**POPPY Study**

**Topic Guide to inform interviews with commissioners**

1. What prompts decision to commission a particular service?

2a. When commissioning services for older adults, what costings/economic evaluation do you normally look for? (e.g., location, staffing, resources, third sector partnerships, community facilities)

Or

2b. When commissioning a pain service, what costings/economic evaluation do you normally look for? (e.g., location, staffing, resources, third sector partnerships, community facilities)

3. How much flexibility do you have with costings/budgets?

3. When commissioning a pain service for older adults what outcomes do/would you consider? For example:

- Policy recommendations.
- Data on population need.
- Patient reported outcomes (e.g., independence, self-care, pain, mental health/mood)
- Clinician reported outcomes (e.g., clinician perceived improvement, number of sessions completed, number of referrals to third sector partners (gym/weight loss/smoking cessation), waiting list times, number of patients attending/completing pain management programmes, impact on care emergency, impact on health care, reduced contacts in primary care, opioid prescribing).
- What would you prioritise to inform commissioning decisions?

4. What staffing levels do you/would you look for and why? For example, expertise, experience, grade of staff, e.g. more expensive clinical psychologists versus less expensive Improving Access to Psychological Therapies (IAPT) therapists?

5. What facilities do you/would you expect to be available and why? For example, clinic space, community halls, gyms, equipment?

6. Do you/would you consider the team's reputation and whether they can deliver the service? What factors would determine whether the team can deliver the service?

7. Are you interested in how the team aim to develop their service/audit/undertake research? What information do/would you expect to see from the team?

8. How do/would you monitor the service once it has been commissioned - what ongoing data do you request from the service providers?

9. What support (if any) do you/would you provide to the service?

10. How do you manage problems/complaints and when do you implement action if services are underperforming?

11. Are there any specific services in your areas that provide support for older people living with frailty and pain, or plans for services for this population?

12. What information would you want to see in a commissioning guide for pain and frailty services?  This is the main planned output for the POPPY study.
