## Supplementary File 3 Case Study for "Maximising pain services for frail older adults, the views of healthcare professionals and commissioners: findings from the Pain in Older People with Frailty (POPPY) study"

### CASE STUDY: Mr Brian Kelly

Brief Intro

81, lives with wife who has dementia in a town in Somerset. He served in the Navy for 30 years until retirement 23 years ago.

General health and wellbeing

- His general health is very good, he has a good diet and healthy lifestyle.
- He had surgery on his lower spine 5-6 years ago due to slipped discs which was effective as was his previous heart surgery several years ago.
- He has been a very sporty man in the past and enjoyed cycling, running and swimming. He gave up long distance cycling only a few years ago, following surgery on his spine. But he walks for about an hour every morning when he takes the dog out and also has a gym set up in his garage where he regularly exercises.

Pain

- He has pain in his shoulders and back, which has been ongoing and undiagnosed for over 30 years. He believes heavy lifting and carrying (without guidance on doing this safely) in the Navy has contributed to his current pain.
- His most recent pain is in his hips which appears to be getting worse. After a physical examination of his hip, a couple of months ago, his GP suggested it could be age-related/arthritis and offered him paracetamol and co-codamol.

Pain impact on daily life

- He enjoys gardening but struggles to bend and lift now.
- He does all the cooking and cleaning but must do things in stages.
- He still drives and goes shopping, which is his ‘me’ time.
- He is organised in his caring role as he is mentally alert but recognises, he is slowing down physically.

Attitude to living with pain

- He is not a moaner and has a very positive attitude- his time in the Navy has helped with his overall approach to life – to stay positive and not give in.
- He does not consider caring for his wife to be a burden as it appears to keep him going and motivates him to ‘fight’ the pain.

Management strategies:

- He uses heat patches which offer some relief and he does stretching exercises on his kitchen floor.
- He tries to avoid taking pain killers, even when prescribed by his GP and has told his GP that he does not want to take Co-codamol.
- He had tried physiotherapy in the past which has not worked for him long-term.
- He would not consider surgery as he needs to be able to care for his wife.

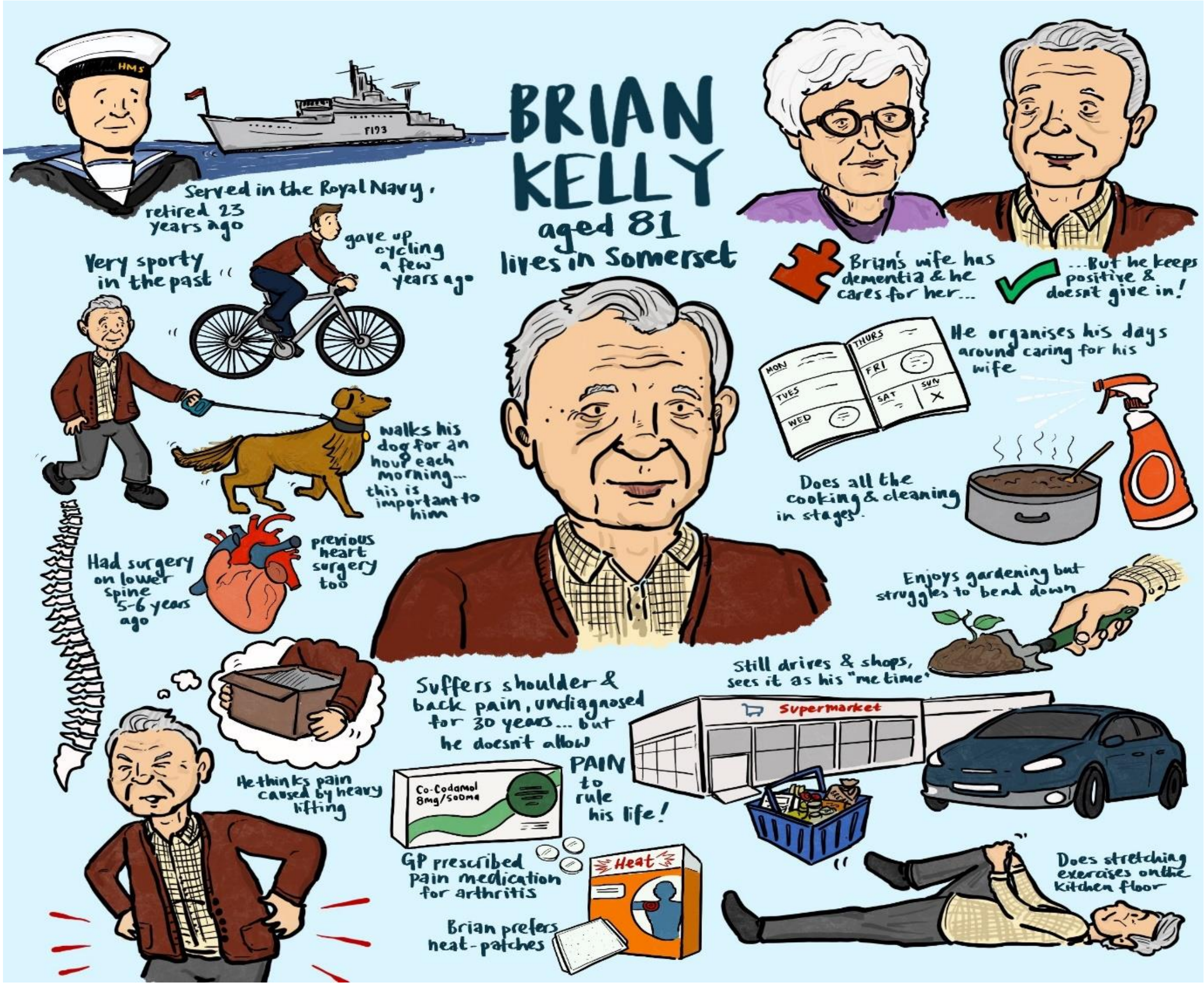
BRIAN KELLY
aged 81
lives in Somerset
Served in the Royal Navy, retired 23 years ago
Very sporty in the past
gave up cycling a few years ago
walks his dog for an hour each morning... this is important to him
Had surgery on lower spine 5-6 years ago
previous heart surgery too
He thinks pain caused by heavy lifting
Brian's wife has dementia & he cares for her...
...But he keeps positive & doesn't give in!
He organises his days around caring for his wife
MON
TUES
WED
THURS
FRI
SAT
SUN
Does all the cooking & cleaning in stages.
Enjoys gardening but struggles to bend down
Suffers shoulder & back pain, undiagnosed for 30 years... but he doesn't allow PAIN to rule his life!
Co-Codamol 8mg/500mg
GP prescribed pain medication for arthritis
Heat
Brian prefers heat-patches
Still drives & shops, sees it as his "me time"
Supermarket
Does stretching exercises on the kitchen floor
