## Supplementary File4 Final Coding Framework for "Maximising pain services for frail older adults, the views of healthcare professionals and commissioners: findings from the Pain in Older People with Frailty (POPPY) study"

### Final Coding Framework
### HCP views and experiences of existing pain management services for older people

#### Design and implementation of pain service

- Commissioning pain services (incl. factors influencing commissioning processes)
- Initial development of service (incl. start-up date)
- Service configuration (incl. location of service; alignment to other services i.e., Tertiary/Hospital based/community)
- Ethos/Aims of pain service
- Staff skill mix /HCP roles in service (incl. relevant prof. background)
- Identification of service users (incl. referral pathway/sharing service information with public/ Screening / triage of service users)
- Service user characteristics (age; frailty; pain status; catchment)
- Service user assessment
- Pre PMP preparation activities
- Identification of service user goals (incl. choice of treatment)
- Service components (elements of PMP/content of sessions/treatment options)
- Frequency and duration of interventions (incl. size of group)
- Format of sessions (inclusion of family/carers; face to face vs online; individual out-patient work)
- Working with other services
- Specific service/approaches to working with OAs (or not)
- Review/disengagement/discharge processes
- Measurement of service user outcomes
- Service evaluation

#### Service User response to service

- Barriers for service user engagement in service
- Barriers to positive outcomes
- Facilitators for service user engagement in service
- Facilitators of positive outcomes
- Age related responses to service

#### HCP/commissioner views/experiences of service(s)

- Contextual factors impacting service provision (socioeconomic/geographic)
- Relevant learning/skills developed during service delivery
- Facilitators of service delivery
- Barriers/ challenges to service delivery
- Flaws/disadvantages of service (including potential threats to service)
- Pain service improvements (suggested/actioned)
- Barriers to service improvements
- Facilitators of service improvements
- Attitudes to pain management in general (incl. views on specialist OA/frailty service)
- Strengths/advantages of pain service
